# Temporal Relationships between Smartphone Application Use and Online Substance Procurement in U.S. Youth

**DOI:** 10.64898/2026.05.15.26353324

**Authors:** Meredith Gansner, Margaret Adams, Prakruthi Nikam, Nyah Huntley, Sai Ramrajesh, Lisa A. Marsch, Sharon Levy, Zev Schuman-Olivier

**Affiliations:** University of Rochester Medical Center, Rochester, NY, USA; Boston Children’s Hospital, Boston, MA, USA; Center for Technology and Behavioral Health, Dartmouth Geisel School of Medicine, Lebanon, NH, USA; Cambridge Health Alliance, Cambridge, MA, USA

## Abstract

**Background:** Despite the significant risks associated with online substance procurement (SP), few researchers have examined this practice in U.S. youth. The studies that do exist are cross-sectional and cannot temporally connect specific digital behaviors to online SP. This longitudinal cohort study examined youth SP and digital media habits to determine whether use of certain smartphone applications correlated with increased odds of online SP or being contacted online about procuring drugs or alcohol.

**Methods:** A cohort of U.S. youth (aged 15-20) with a history of non-daily substance use in the 3 months prior to enrollment was recruited to use the digital phenotyping smartphone application EARS for 90 days. On a nightly basis, participants were asked to complete surveys about online experiences related to SP and instances of substance use. Smartphone-generated screen use data were also collected passively each day.

**Results:** Out of 112 enrolled participants, 106 were able to be included in analyses. Over approximately 3 months, 28.3% of participants (n=30) reported a collective 91 instances where they used social media to acquire drugs or alcohol. Screen use data demonstrated temporal relationships between social media SP and applications previously connected to the social media drug-purchasing process (e.g., TikTok, encrypted apps), as well as other school-specific social media.

**Discussion:** Our results provide critically needed research evidence to support a body of literature composed predominantly of anecdotal reports. Despite measures taken by social media companies to prevent use of their platforms for drug procurement, underage youth continue to engage in this practice.

## INTRODUCTION

The recent epidemic of overdose deaths among youth in the United States has been attributed to illicit prescription drugs being laced with fentanyl.^1–3^ While challenging to confirm through national datasets, multiple investigative reports performed by the lay media have highlighted the role of social media platforms in facilitating youth access to contaminated pills.^4–8^ Existing research confirms that social media streamline the marketing and selling of illicit substances through their ability to disseminate product information rapidly to a wide audience.^9–16^ For underage youth, social media substance procurement (SMSP) may appear especially enticing because it allows for covert and expedient access to specific substances via familiar technologies.^16,17^

Instances of SMSP by adolescents and young adults may represent age-typical risk-taking behaviors involving recreational substance use and social gatherings featuring sharing of drugs and alcohol, now facilitated through social media-based communication. For example, an adolescent’s friends could message them through a social media platform, offering to share acquired drugs or alcohol, or informing them of a locale where those substances could be acquired. However, a 2022 report published by the U.S. Drug Enforcement Administration (DEA) also cautions about the existence of a virtual “marketplace” for drugs and alcohol rooted on popular social media platforms, established by illicit drug traffickers.^18^ The report outlines these online transactions as a three-stage process where sellers first advertise a product on mainstream social media platforms using coded language or emojis to evade detection by platform censors. Interested buyers then initiate contact with the seller through public posting or direct messaging, and further communication transitions to more secure encrypted messaging apps, with payment remitted through cash exchange applications like Zelle or Venmo.^18^

Little research has examined SMSP by adolescent and young adult cohorts despite the significant risks. In a survey of U.S. individuals aged 15-25, 2% of all respondents reported using online sources to buy drugs, with over half of the purchases made via social media.^19^ Another study based in the United Kingdom reported that 10% of their adolescent participants had purchased drugs through social media, and that Snapchat, text messaging, and Instagram were the apps most commonly used to facilitate these transactions.^20^ While well-powered, these cross-sectional studies have notable limitations. By focusing on incidence rates within the overall adolescent population, researchers are unable to collect detailed information about online SP within higher-risk groups, such as those with a current or recent history of substance use. Furthermore, results from cross-sectional survey studies are likely impacted by recall bias and unable to offer substantive evidence of significant relationships between certain types of digital media use and online drug procurement.

Social media content analyses have provided detailed information about what drug-purchasing content is easily accessible to underage youth on specific social media platforms. Compiling results of these studies, it has been approximated that every 13 of 100 social media posts are related to drug and alcohol procurement.^21^ However, this methodology cannot temporally connect use of specific online platforms to instances of online drug procurement. Therefore, relatively little is known about the granular mechanisms used by youth to procure drugs and alcohol in their online environment.

The current study examined online SP in U.S. youth using a smartphone-based ecological momentary assessment (EMA) protocol. Using both daily self-report surveys and data collected passively through smartphone sensors, the study investigated whether use of specific smartphone applications correlated with increased odds of 1) SP (any) and SMSP and 2) being contacted via digital media about obtaining drugs or alcohol both via direct message/text (DM/text contact) and social media/online (SM/online contact). Lastly, this study also explored whether a temporal relationship exists between the proportion of negative words participants use in typed content and the likelihood of DM/text contact and SM/online contact. Peers who have access to substances may be more likely to message a friend with an offer of drugs or alcohol if that friend’s posts suggest psychological distress. And, while not yet confirmed in the research literature, there is some evidence that professional drug sellers may proactively target potential adolescent buyers, initiating contact with youth they perceive as more vulnerable.^22^

Given their established role in facilitating drug-related communications, as well as their capacity to provide enhanced anonymity and privacy in digital exchanges, we hypothesized: 1) odds of using encrypted messaging applications and platforms featuring 24-hour automatic message deletion (e.g., Snapchat) would be significantly higher on days when SMSP took place, and 2) days where youth reported DM/text contact and SM/online contact would feature a greater percentage of typed words indicating a negative mood/affect.

## METHODS

### Study Design

This longitudinal cohort study examined SP, substance use, and digital media habits in a cohort of U.S. youth between the ages of 15 and 20 over 90 days using the digital phenotyping smartphone application EARS to collect daily surveys and passive sensor-derived data. Data were collected between August 2024 and August 2025. The study protocol was approved by the hospital system’s institutional review board.

### Recruitment

Potential participants were between the ages of 15 and 20 at the time of enrollment, lived in the United States, owned a personal smartphone (Android or iPhone), and used predominantly English when typing on their smartphones. Participants were not eligible to participate in the study if they reported substance use every day in the 3 months prior to enrollment.

Prospective participants were recruited nationally through multiple methods, including social media advertising via Facebook and Instagram, distributing study flyers to national listservs, posting flyers near high school and college campuses around the United States, and hosting informational recruitment tables at local adolescent health clinics. Study flyers featured a QR code that linked to a REDCap survey where interested youth could enter their cell phone number to be contacted by the study team. When the study team received a youth’s cell phone number, the PI sent a text message using the secure health application Doximity and a follow-up phone call was arranged to discuss the study in greater detail and perform an eligibility screen.

If a potential participant was 18 years of age or older and met study eligibility criteria, a video meeting was scheduled to complete the informed consent process and help the participant install and set up the EARS application on their smartphone. If a potential participant was eligible but under 18 years of age, a subsequent video meeting was scheduled for when a parent/guardian was available so that the research team could discuss the study with the parent/guardian and proceed to informed consent and enrollment if the parent/guardian gave their consent for their child to participate. Assent from the participant was also documented during this video meeting.

At the time of study enrollment, participant age, sex, and race/ethnicity were also collected.

### Data Collection and Participant Compensation

Participants used the smartphone-based digital phenotyping application EARS (created by Ksana Health) to complete daily surveys for 90 days. Study participants were sent nightly push notifications at 8:00pm to remind them to complete the daily surveys; each day’s surveys only remained available for completion until midnight the following day. Surveys asked about procurement of drugs and alcohol and instances of digital communication regarding the procurement of drugs or alcohol. The exact wording of these four questions can be found in the supplementary material (**Supplementary Table 1**). Questions asked about “purchasing” *and* “obtaining” through social media to allow for instances of participants procuring drugs or alcohol outside of a financial transaction (e.g., given for free or bartered). Youth were also asked each day about intentional engagement with substance-related digital content: posting on social media about drugs or alcohol, sending direct messages about drugs or alcohol, and searching for articles or forums related to drugs or alcohol.

Because prior research has pointed to youth use of text messaging to procure substances,^20^ and text messaging applications are not considered social media, the study questionnaire asked about online procurement-related contact both via direct message/text and social media platform specifically. Youth were not asked to disclose the identity of the person who contacted them about acquiring drugs or alcohol. Information about daily app use was collected in two separate ways depending upon the participant’s smartphone operating system. For Android-using participants, app use data were collected passively without any additional action required on the part of the participant. For iPhone-using participants, app use data generated through iOS were uploaded into the EARS application each night at the time of the active surveys. The EARS application would then prompt participants to take screenshots of uploaded app use data, which were automatically uploaded into the EARS app. Typed content was recorded passively through the EARS app’s keyboard logger. Only content typed using the EARS keyboard was examined. Based on the analytic approach used by Byrne et al. to explore relationships between affective language of typed content and stress,^23^ Ksana Health’s data science team can calculate the proportion of words associated with negative affect that were typed into the EARS keyboard on a given day.^24^ Our study team received only the proportion of words typed daily indicating positive or negative affect, no raw keyboard data. All collected data were downloaded from participant phones and uploaded to a secure AWS server accessible only to study team members.

Participants were compensated up to $250 via ClinCard for their participation, a fixed $35 at the beginning and end of the study period, and between $10 and $20 each 10-day period dependent on percentage of daily surveys completed during those 10 days (5 or more completed daily surveys in a 10-day period: $20, < 5 completed daily surveys in a 10-day period: $10).

### Data Processing & Analysis

Raw data were processed using Excel. App use was coded either “1” (used that day) or “0” (did not use that day). Based on the mechanism for social media drug purchasing outlined in the U.S. Drug Enforcement Administration’s drug trafficking threat report,^18^ only applications with direct messaging capabilities or those previously implicated in drug procurement were included in analyses. Answers from two EMA survey questions were used to generate two additional outcomes of interest regarding instances of unsolicited online procurement-related contact: 1) instances where a participant reported SM/online contact *without* the participant reporting same-day substance-related online activities, and 2) instances where a participant reported DM/text contact *without* the participant reporting same-day substance-related online activities. Because a participant’s active engagement with drug and alcohol-related online content could theoretically influence their being contacted about purchasing substances (e.g., “liking” a social media post that depicts cannabis use prompts messaging from a drug seller), online contact regarding buying drugs or alcohol was *only considered unsolicited* on days when the participant: 1) did not report procuring substances through social media, 2) did not report posting on social media about drugs or alcohol, 3) did not report sending direct messages to others about drugs or alcohol, and 4) did not report searching for articles or forums related to drugs or alcohol. To ensure adequate power for analyses, encrypted messaging apps (i.e., WhatsApp, Telegram, and Signal) and food service apps (i.e., UberEats, DoorDash, and GrubHub) were combined into single categories. Only apps used by 5 or more participants were included.

Analyses were performed using SPSS 31.0 and R 4.5.2. Mixed-effects logistic regression models were used to examine associations between app use and the following outcomes: SP (any), SMSP, SM/online contact, and DM/text contact. Methods for handling missing data were tailored to data structure under a missing-at-random assumption. For SP variables and online procurement-related contact variables (both binary outcome variables), missing data were addressed using multiple imputation implemented via mice. Given sparsity in binary outcomes and predictors, a single-level imputation approach was used, with variables imputed using logistic regression models. Twenty imputed datasets were generated, and each was analyzed using generalized linear mixed-effects models with a logit link and random intercepts to account for clustering within individuals, implemented in lme4. Parameter estimates were pooled using Rubin’s rules. A full information maximum likelihood (FIML) model was used to examine the association between the proportion of negative words typed and instances of unsolicited, one-way online procurement-related contact via SM/online contact or via DM/text contact. Statistical precision was determined with 95% confidence intervals. To account for multiple comparisons, p-values were adjusted using the Benjamini–Hochberg false discovery rate (FDR) procedure, and statistical significance was evaluated using the FDR-adjusted p-values.

## RESULTS

A total of 112 participants enrolled in the study, and data from 106 individuals were available for the analyses. Of the 6 individuals who were excluded from analyses, 3 were lost to follow-up before contributing any survey data and 3 were later found to be ineligible due to age or location. The sample ranged in age ranged from 15 to 20 years, with a mean age of 18.6 (SD: 1.1) (**Table 1**). The sample was of predominantly female sex (n=72, 67.9%), white non-Hispanic/Latine race and ethnicity (n=54, 50.9%) (**Table 1**).

**Table 1:** Sample Demographics, Substance-Related Contact and Procurement, and Substance Use Patterns (N=106)

| Demographics |  |  |
| --- | --- | --- |
| Mean Age (SD) | 18.6 (1.1) |  |
| Sex, n (%) |  |  |
|  | Female | 72 (67.9) |
|  | Male | 31 (29.2) |
|  | Did Not Answer | 3 (2.8) |
| Race/Ethnicity, n (%) |  |  |
|  | White non-Hispanic/Latine | 54 (50.9) |
|  | <sup>a</sup>Racial/Ethnic Minority | 39 (36.8) |
|  | *Black* | 12 (30.8) |
|  | *Asian* | 10 (25.6) |
|  | *Multiracial* | 9 (23.1) |
|  | *Hispanic/Latine* | 8 (20.5) |
|  | *Did Not Answer* | 13 (12.3) |
| Substance Procurement-Related Contact |  |  |
| Contact Type | Instances: n | Participants: n (%) |
| Contacted via DM/Text | 445 | 73 (68.9) |
| Contacted Online or on SM | 357 | 69 (65.1) |
| Contacted Online or on SM *without* same-day DM/Text Contact | 120 | 40 (37.7) |
| Substance Procurement |  |  |
|  | Instances: n | Participants: n (%) |
| Any Substance Procurement | 769 | 92 (86.8) |
| SMSP<sup>b</sup> | 91 | 30 (28.3) |
| Substance Use Reported on Days with SMSP |  |  |
| Substance Type | + SMSP: n (%) | - SMSP: n (%) |
| Cannabis | 62 (16.4) | 316 (83.6) |
| Tobacco/Nicotine | 46 (26.0) | 131 (74.0) |
| Alcohol | 45 (9.6) | 425 (90.4) |
| Prescription Drug | 8 (40.0) | 12 (60.0) |
| Illegal Drug | 2 (28.6) | 5 (71.4) |

SMSP: social media substance procurement
<sup>a</sup>All participants who do not identify as White, non-Hispanic/Latine.
Racial/ethnic minority subgroup percentages: denominator is total number of participants who identified as a racial/ethnic minority (n=39)

There were 769 reports of SP (any), representing 86.8% of total participants (n=92). Of these 769 instances, only 91 (11.8%) were obtained through SMSP (**Table 1**). However, out of the 92 participants who obtained drugs or alcohol at some point during the study, 30 (32.6%) did so via social media at least once during the 90-day study period (28.3% of the overall 106 participants). Cannabis was the substance used most frequently on days when SMSP was reported (n=62, 68.1%), followed by nicotine (n=46, 50.5%) and alcohol (n=45, 49.5%). However, when examining how often SMSP occurred on the same day as substance use, non-medical prescription drug use had the highest same-day co-occurrence with SMSP, with SMSP reported on 40.0% (8/20) of use days, followed by illegal drug use at 28.6% (2/7) (**Table 1**). Across the study period, 68.9% of participants reported at least one instance of DM/text contact (445 total instances). Similarly, 65.1% of participants reported at least one instance of SM/online contact (357 total instances). Of the 357 instances where participants reported SM/online contact, 33.6% were isolated to SM/online contact and were not accompanied by same-day DM/text contact (**Table 1**).

On days when study participants used encrypted apps, TikTok, and food delivery apps, they had significantly higher odds of SP (any) (Encrypted apps: OR=1.74, 95% CI:1.25-2.43, p=0.01; TikTok: OR=1.81, 95% CI:1.29-2.54, p=0.01; Food delivery apps: OR=1.81, 95% CI:1.26-2.61, p=0.01) (**Figure 1**). There were significantly higher odds of SMSP on days when encrypted messaging apps and Fizz, an anonymous social media platform for high school and college students, were used (Encrypted apps: OR=2.85, 95% CI:1.56-5.22, p=0.02; Fizz: OR=4.70, 95% CI:1.94-11.37, p=0.02) (**Figure 1**).

**Figure 1.**
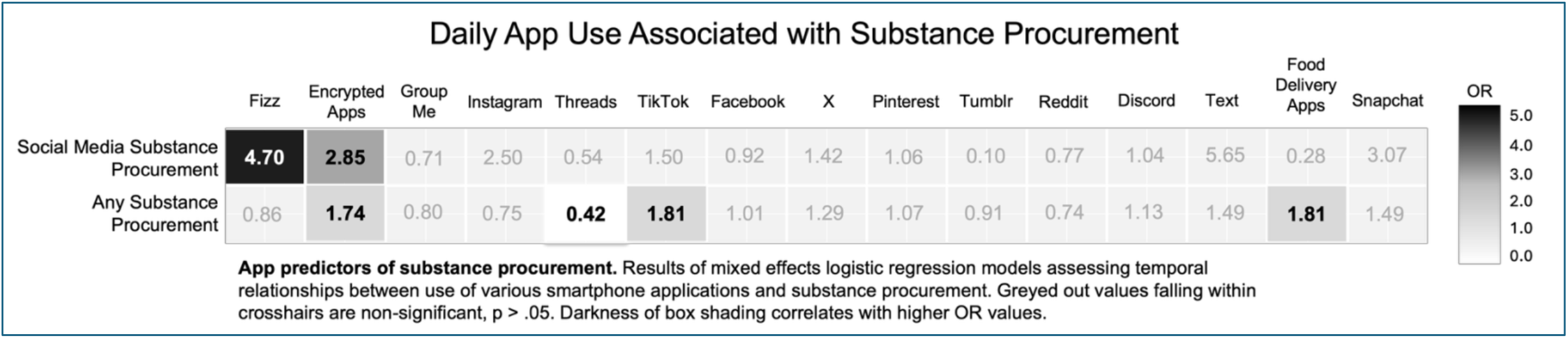

Similar patterns of app use were seen when examining digital contact received from others regarding purchasing or accessing drugs or alcohol. The odds of receiving DM/text contact were significantly higher on days when TikTok was used (OR=2.23, 95% CI:1.38-3.63, p=0.03) (**Figure 2**). The odds of receiving SM/online contact were significantly higher on days when TikTok and encrypted apps were used (TikTok: OR=2.78, 95% CI:1.69-4.58, p=0.001; Encrypted apps: OR=2.89, 95% CI:1.89-4.42, p<0.001) (**Figure 2**).

**Figure 2.**
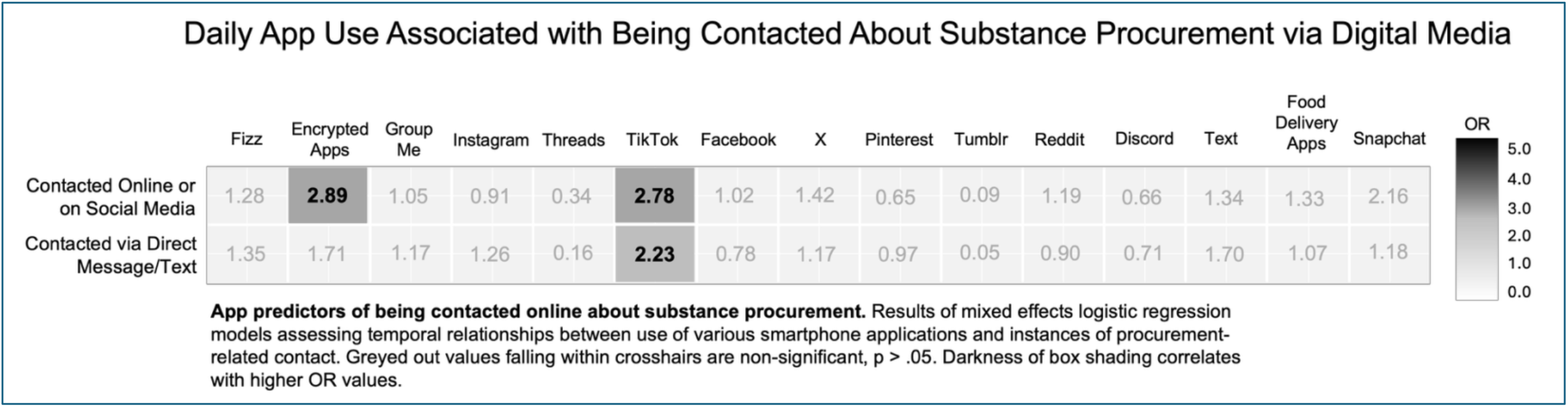

A significant relationship was also found between the sentiment of a participant’s typed content and being contacted via digital media about substance acquisition; however, the positive correlation was only seen when the contact occurred through social media (SM/online: β=0.22, SE: 0.09, p=0.02; DM/text: β= -0.07, SE: 0.08, p=0.35). Even on days when participants reported no substance-related messaging or engagement with any substance-related online content, the significant association remained (SM/online: β=0.54, SE: 0.16, p=0.001; DM/text: β= -0.18, SE: 0.14, p=0.21).

## DISCUSSION

To our knowledge, we are the first to study SMSP in a cohort of U.S. youth using a combination of ecological momentary assessment surveys and app use data. In just 3 months, 30 underage youth collectively reported 91 instances where they used social media to acquire drugs or alcohol; their screen use data demonstrated temporal relationships between applications previously connected to the social media drug-purchasing process (e.g., TikTok, encrypted apps), as well as other school-specific social media. Our results highlight several notable findings concerning online SP in this demographic: 1) SMSP may be more common among youth than previously reported, 2) SMSP includes use of school-specific social media for youth affiliated with academic institutions, 3) expressing negative sentiment online may increase the likelihood that an adolescent is contacted via social media about substance procurement, and 4) significant temporal relationships between SMSP, online procurement-related contact, and use of both TikTok and encrypted apps suggest youth continue to engage in SMSP via the specific multi-step process outlined in the 2022 U.S. DEA report.^18^

### Prevalence of Social Media Drug Acquisition

Prior cross-sectional studies examining online SP across overall samples of youth found prevalence rates ranging from 2.0% to 10.0%.^19,20^ Even among youth with a history of drug and alcohol use, rates as high as ours have not been reported. Oksanen et al. found that prevalence increased from 2.0% to 10.0% when only including participants with a past history of non-alcohol/tobacco drug use.^19^ One cohort of adolescent and young adult patients in substance use treatment programs found that only 2.3% of respondents had purchased prescription pills online.^15^

Our SMSP prevalence rate of 28.3% could potentially be explained by differences in both methodology and timing. Answering questions on personal smartphones in a familiar setting, EMA study participants may feel more comfortable answering questions about their high-risk or illegal behaviors, including drug use.^25^ Additionally, our study asked about all substances, and not just substance purchasing but any procurement, encompassing a wider range of transactions. Lastly, most prior research studying youth online SP was published before 2020. Drug use in high school-aged populations has been declining in the United States,^26^ and social gatherings where youth may have previously experimented with drugs and alcohol were limited during the COVID-19 pandemic,^27^ when many of our study participants were in high school. Decreased exposure to offline methods of SP *and* excessive time spent using digital media may have helped SMSP become increasingly accepted in this demographic within the last four to five years.

Our findings related to types of substance use associated with SMSP were similar to prior studies. Cannabis has previously been found to be the drug most frequently purchased on social media,^16,17,28,29^ and in our cohort, cannabis was the drug used most on days when SMSP was reported. However, the likelihood of an individual using social media to procure a drug may also be drug dependent. The 2025 Monitoring the Future survey found that between 2020 and 2024, only 3.4% of 12^th^ graders who used amphetamines bought them online, compared to 13.3% of the 12^th^ graders who had used non-heroin narcotics.^26^ Few instances of prescription drug misuse were reported by our participants (n=20), but when they did occur, 40.0% of them accompanied SMSP (compared to only 16.4% of total instances of cannabis use). Less commonly used substances (e.g., prescription opioids) are likely harder for youth to access, and the convenience of social media purchasing may help youth justify SMSP-associated risks, like fentanyl contamination.

### Platform Use Associated with Substance Procurement and Related Communications

Several platforms linked to both SMSP and online procurement-related contact matched our hypotheses, as well as descriptions of the social media purchasing process in the United States and other countries.^18,30^ Despite estimates that nearly 68% of U.S. teenagers use TikTok^31^ and 16.0% report “almost constant” use,^32^ we were able to detect a significant difference in odds of TikTok use on days when online procurement-related contact was reported. Because of its popularity, TikTok maximizes the number of individuals exposed to drug sellers’ advertising, and direct messaging and video commentary^30^ offer opportunities for instantaneous connection to a local dealer. Similarly, because encrypted apps are reportedly where most seller/buyer communication occurs (presumably to protect seller anonymity), this may explain the significant relationship between encrypted app use and all four of the substance-related outcomes we examined. The relationship between SP and encrypted app use was strongest for SMSP specifically (OR=2.85 vs. OR=1.74), but it may be that some underage youth use encrypted messaging apps to discuss any procurement of drugs or alcohol, even with known acquaintances, like friends or classmates.

A couple of significant relationships between app use and drug-related outcomes were not anticipated, specifically Fizz and food-delivery platforms. To our knowledge, Fizz has not previously been recognized for its role in drug-related transactions in either lay media reports or research literature. Like most online forums, there are platform guidelines citing acceptable content, but moderation is the responsibility of student members^33^ and guidelines may not be consistently enforced. However, because the app does require users to verify their cellphone number and school e-mail address, adolescents may perceive Fizz as a comparatively safer method of drug procurement. Future studies are needed to clarify how anonymous school-based apps connect students to substances. While food delivery services could theoretically enable underage youth to access cannabis in states where it is legalized (e.g., driver doesn’t verify age of the buyer), cannabis and THC delivery is highly restricted, with DoorDash alone delivering only hemp-derived THC and CBD products.^34,35^ Therefore, rather than representing a means for procurement, it appears more likely that youth are ordering food through these applications when they use substances.

Because our participants were not asked names of the specific apps they used to obtain substances, nor names of apps on which they were messaged/contacted, these findings must not be interpreted as proof of causal relationships. However, objective measures of daily app use increase confidence that platforms like TikTok and Fizz, as well as encrypted apps like Signal and WhatsApp, are in some way relevant to youths’ online substance acquisition. For example, while our results connecting TikTok and online procurement-related contact support prior evidence of the platform’s role in initiating contact with online drug sellers,^18,36^ TikTok’s influence on SP could also manifest in other ways. Lay media articles highlight a growing #Pingtok community where youth post videos of themselves intoxicated and seek information about accessing drugs.^30^ Thus, the temporal link between TikTok use and online procurement-related contact could also be the result of youth being triggered to procure substances after exposure to drug-related TikTok content.

### Sentiment of Typed Content and Unsolicited Online Contact about Substance Acquisition

Because negative emotions are well-established drivers of substance use,^37,38^ it was expected that participants’ typed content would display more negative affect on days when they were contacted about procuring drugs or alcohol. Frequently, instances of being contacted about SP were accompanied by reports of same-day substance-related messaging initiated by the participant. However, it is concerning that even when participants did not endorse substance-related messaging, there was a significant relationship between affective content of typed words and receiving online procurement-related contact. Because both expression of negative emotions on social media and engagement with mental health-related posts have been shown to prompt algorithmic recommendations of harmful content (e.g., drug use, suicide, self-harm),^22,39,40^ it will be important to determine both the creators of, and language used in, these procurement-related social media contacts.

### Limitations

While better able to capture the nuances of temporal relationships, protocols using smartphone-based EMA have their own limitations. Particularly when studying high-risk adolescent behaviors, responsible conduct of research requires prioritization of participant safety with repeated monitoring and check-ins, often at the expense of a large sample size. Thus, our sample of 106 youth is a national convenience sample and therefore not nationally representative. Future studies should focus on optimizing inclusion of young males, particularly as they are overrepresented in substance-related deaths.^41^

Additionally, EMA surveys must balance a need for continued participant engagement with a need to gather pertinent information through active survey questions. We employed passive data collection as much as was possible and relevant for the study, but an inability to access precise details of participants’ online experiences still leaves many questions about SMSP unanswered. While their use is presently restricted to select smartphone operating systems, programs that enable passive collection of screenshot data may be one way to mitigate this issue in subsequent research.

### Conclusion

Despite measures taken by social media companies to prevent use of their platforms for drug procurement, this study demonstrates that SMSP continues to occur among underage youth in the United States. In particular, adolescents may be more likely to acquire certain substances, like prescription pills, through SMSP. Our findings highlight a key concern for parents, clinicians, educators, and policymakers: by reducing access barriers, SMSP may increase opportunities for reactive, solitary substance use, underscoring the need for close monitoring of substance use patterns among older adolescents and young adults as unrestricted access could reshape how drug and alcohol experimentation occurs during this developmental period.

## Data Availability

All data produced in the present study are available upon reasonable request to the authors

---

**SUPPLEMENTARY TABLE 1:** OUTCOME QUESTIONS.

**Figure 2.**
| SUPPLEMENTARY TABLE 1: OUTCOME QUESTIONS |
| --- |
| **1). Contact via social media about procuring substances** |
| TODAY, did anyone contact you ONLINE or over SOCIAL MEDIA about purchasing or accessing drugs or alcohol?<br>• Yes<br>• No |
| **2) Contact via direct message about procuring substances** |
| TODAY, did anyone contact you through a DIRECT MESSAGE or TEXT about purchasing or accessing drugs or alcohol?<br>• Yes<br>• No |
| **3) Obtained substances that day (any method of procurement)** |
| TODAY, did you obtain drugs or alcohol in any way?<br>• Yes<br>• No |
| **4) Obtained substances that day using social media** |
| If yes to question #3:<br>TODAY, did you purchase or obtain drugs or alcohol using a social media platform?<br>• Yes<br>• No |

## References

1. Friedman J, Godvin M, Shover CL, Gone JP, Hansen H, Schriger DL. Trends in Drug Overdose Deaths Among US Adolescents, January 2010 to June 2021. JAMA. 2022;327(14):1398. doi:10.1001/jama.2022.2847

2. Tanz LJ, Dinwiddie AT, Mattson CL, O’Donnell J, Davis NL. Drug Overdose Deaths Among Persons Aged 10–19 Years — United States, July 2019–December 2021. MMWR Morb Mortal Wkly Rep. 2022;71. doi:10.15585/mmwr.mm7150a2

3. O’Donnell J, Tanz LJ, Gladden RM, Davis NL, Bitting J. Trends in and Characteristics of Drug Overdose Deaths Involving Illicitly Manufactured Fentanyls — United States, 2019–2020. MMWR Morb Mortal Wkly Rep. 2021;70. doi:10.15585/mmwr.mm7050e3

4. Ortutay B. How social media became a storefront for deadly fake pills laced with fentanyl. PBS NewsHour. September 12, 2024. Accessed April 16, 2026. https://www.pbs.org/newshour/nation/how-social-media-became-a-storefront-for-deadly-fake-pills-laced-with-fentanyl

5. Mann B. Social media platforms face pressure to stop online drug dealers who target kids. NPR. January 26, 2023. Accessed April 16, 2026. https://www.npr.org/2023/01/26/1151474285/social-media-platforms-face-pressure-to-stop-online-drug-dealers-who-target-kids

6. Organization for Social Media Safety. Social Media & Drugs: What Parents Need To Know. Organization for Social Media Safety. January 12, 2022. Accessed April 16, 2026. https://www.socialmediasafety.org/blog/social-media-drugs-what-parents-need-to-know/

7. Hoffman J. Fentanyl-Tainted Pills Bought on Social Media Cause Youth Drug Deaths to Soar. N Y Times. May 19, 2022. Accessed April 16, 2026. https://www.nytimes.com/2022/05/19/health/pills-fentanyl-social-media.html

8. Drug Enforcement Administration. Sharp Increase in Fake Prescription Pills Containing Fentanyl and Meth. Drug Enforcement Administration; 2021:1–2. Accessed April 16, 2026. https://www.dea.gov/alert/sharp-increase-fake-prescription-pills-containing-fentanyl-and-meth

9. Hu C, Yi M, Liu B, Li X, Ye Y. Detection of Illicit Drug Trafficking Events on Instagram: A Deep Multimodal Multilabel Learning Approach. Proc 30th ACM Int’l Conf Inf Knowl Manag CIKM ‘21 Novemb 1–5 2021 Virtual Event Aust. Published online October 2021:3838–3846. doi:10.1145/3459637.3481908

10. Kong G, Schott AS, Lee J, Dashtian H, Murthy D. Understanding e-cigarette content and promotion on YouTube through machine learning. Tob Control. 2023;32(6). doi: 10.1136/tobaccocontrol-2021-057243

11. Crocetti A, Lister N, Martino F, et al. Alcohol Promotion via User-Generated Content on Instagram and TikTok: A Content Analysis. Drug Alcohol Rev. 2025;44(7):2019–2029. doi: 10.1111/dar.70027

12. Whitehill JM, Trangenstein PJ, Jenkins MC, Jernigan DH, Moreno MA. Exposure to Cannabis Marketing in Social and Traditional Media and Past-Year Use Among Adolescents in States With Legal Retail Cannabis. J Adolesc Health. 2020;66(2):247–254. doi: 10.1016/j.jadohealth.2019.08.024

13. Demant J, Anderdal Bakken S, Oksanen A, Gunnlaugsson H. Drug dealing on Facebook, Snapchat and Instagram: A qualitative analysis of novel drug markets in the Nordic countries. Drug Alcohol Rev. 2019;38:377–385. doi: 10.1111/dar.12932

14. Demant J, Aagesen KMB. An analysis of drug dealing via social media. Eur Monit Cent Drugs Drug Addict. Published online 2022.

15. Festinger DS, Dugosh KL, Clements N, et al. Use of the internet to obtain drugs without a prescription among treatment-involved adolescents and young adults. J Child Adolesc Subst Abuse. 2016;25(5):480–486. doi:10.1080/1067828X.2015.1103345

16. Moyle L, Childs A, Coomber R, Barratt MJ. #Drugsforsale: An exploration of the use of social media and encrypted messaging apps to supply and access drugs. Int J Drug Policy. 2019;63:101–110. doi: 10.1016/j.drugpo.2018.08.005

17. van der Sanden R, Wilkins C, Romeo JS, Rychert M, Barratt MJ. Predictors of using social media to purchase drugs in New Zealand: Findings from a large-scale online survey. Int J Drug Policy. 2021;98. doi: 10.1016/j.drugpo.2021.103430

18. Drug Enforcement Administration. Social Media Drug Trafficking Threat. Drug Enforcement Administration; 2022:1–3. https://www.dea.gov/sites/default/files/2022-03/20220208-DEA_Social%20Media%20Drug%20Trafficking%20Threat%20Overview.pdf

19. Oksanen A, Miller BL, Savolainen I, et al. Illicit drug purchases via social media among American young people. Soc Comput Soc Media. 2020;12194:278–288. doi: 10.1007/978-3-030-49570-1_19

20. Fuller A, Vasek M, Mariconti E, Johnson SD. Platforms, risk perceptions, and reporting: the impact of illicit drug advertisements on social media among UK secondary students. Harm Reduct J. 2025;22. doi:Study Type

21. Fuller A, Vasek M, Mariconti E, Johnson SD. Understanding and preventing the advertisement and sale of illicit drugs to young people through social media: A multidisciplinary scoping review. Drug Alcohol Rev. 2024;43(1):56–74. doi: 10.1111/dar.13716

22. Hanson T. Teens have easier access to drugs as illegal trade booms on social media. CBS News. November 30, 2021. Accessed May 6, 2026. https://www.cbsnews.com/news/social-media-teens-drug-access/

23. Byrne ML, Lind MN, Horn SR, et al. Using mobile sensing data to assess stress: Associations with perceived and lifetime stress, mental health, sleep, and inflammation. Digit Health. 2021;7:20552076211037227. doi:10.1177/20552076211037227

24. Lind MN, Kahn LE, Crowley R, Reed W, Wicks G, Allen NB. Reintroducing the Effortless Assessment Research System (EARS). JMIR Ment Health. 2023;10:e38920. doi: 10.2196/38920

25. Roth AM, Felsher M, Reed M, et al. Potential benefits of using ecological momentary assessment to study high-risk polydrug use. mHealth. 2017;3:46. doi:10.21037/mhealth.2017.10.01

26. Miech RA, Johnston LD, Patrick ME, O’Malley PM. Monitoring the Future Study Annual Report National Survey Results on Drug Use, 1975-2024: Overview and Detailed Results for Secondary School Students. Institute for Social Research, University of Michigan; 2025. Accessed May 7, 2026. https://monitoringthefuture.org/wp-content/uploads/2024/12/mtf2025.pdf

27. Layman HM, Thorisdottir IE, Halldorsdottir T, Sigfusdottir ID, Allegrante JP, Kristjansson AL. Substance Use Among Youth During the COVID-19 Pandemic: a Systematic Review. Curr Psychiatry Rep. 2022;24(6):307–324. doi:10.1007/s11920-022-01338-z

28. van der Sanden R, Wilkins C, Parker K, Rychert M. What app? Demographic and drug use predictors of buying drugs via different social media and messaging apps. Int J Drug Policy. 2026;151:105220. doi:10.1016/j.drugpo.2026.105220

29. van der Sanden R, Wilkins C, Rychert M. “I straight up criminalized myself on messenger”: law enforcement risk management among people who buy and sell drugs on social media. Drugs Educ Prev Policy. 2024;31(3):378–390. doi:10.1080/09687637.2023.2224497

30. Hinz E. TikTok: Teens turn drug use into content. Dtsch Welle. Published online February 7, 2026. Accessed May 8, 2026. https://www.dw.com/en/tiktok-teens-turn-drug-use-into-content-pingtok/a-75807703

31. Faverio M, Sidoti O. Teens, Social Media and AI Chatbots 2025. Pew Research Center; 2025. Accessed May 8, 2026. https://www.pewresearch.org/internet/2025/12/09/teens-social-media-and-ai-chatbots-2025/

32. Faverio M. 10 Facts about Teens and Social Media. Pew Research Center; 2025. Accessed May 8, 2026. https://www.pewresearch.org/short-reads/2025/07/10/10-facts-about-teens-and-social-media/

33. Protect Young Eyes. Fizz App Review. Accessed May 8, 2026. https://www.protectyoungeyes.com/apps/fizz-app-review

34. DoorDash. DoorDash Expands Offerings to Include Hemp-Derived Products in Select States. January 9, 2025. Accessed May 8, 2026. https://about.doordash.com/en-us/news/doordash-launches-hemp-derived-category

35. Stevens B. DoorDash Begins Delivering CBD and Intoxicating Hemp Products. Business of Cannabis. January 15, 2025. Accessed May 8, 2026. https://businessofcannabis.com/doordash-begins-delivering-cbd-and-intoxicating-hemp-products/

36. Dobbs PD, Schisler ED, McCormick C. #Discreetshipping: Selling E-cigarettes on TikTok. Nicotine Tob Res. 2025;27(4):748–752. doi: 10.1093/ntr/ntae081

37. Preston KL, Kowalczyk WJ, Phillips KA, et al. Before and after: craving, mood, and background stress in the hours surrounding drug use and stressful events in patients with opioid-use disorder. Psychopharmacology (Berl). 2018;235(9):2713–2723. doi:10.1007/s00213-018-4966-9

38. Waddell JT, Sher KJ, Piasecki TM. Coping Motives and Negative Affect: An Ecological Study of the Antecedents of Alcohol Craving and Alcohol Use. Psychol Addict Behav. 2021;35(5):565–576. doi:10.1037/adb0000696

39. Molly Rose Foundation. Pervasive-by-Design: TikTok and Instagram. Molly Rose Foundation; 2025:1–40. https://mollyrosefoundation.org/wp-content/uploads/2025/08/proof3_PervasivebyDesign.pdf

40. Amnesty International. Driven Into the Darkness: How TikTok’s “For You” Feed Encourages Self-Harm and Suicidal Ideation. 2023:1–86. Accessed May 9, 2026. https://www.amnesty.org/en/documents/POL40/7350/2023/en/

41. Chen H, Liu S, Wang W, et al. Global burden of substance use disorders in adolescents and young adults aged 10–24 years from 1990 to 2021. Sci Rep. 2025;15(1):25971. doi:10.1038/s41598-025-11266-6

